# Comparing cutaneous NO-dependent vasodilation between young males and females

**DOI:** 10.64898/2026.07.02.26357121

**Authors:** Madison G. Evering, Kelsey S. Schwartz, Claire E. Goebel, Anna E. Stanhewicz, Jody L. Greaney

## Abstract

**Background:** Despite the common use of local heating and intradermal microdialysis perfusion of acetylcholine (ACh) to probe cutaneous endothelium and nitric oxide (NO)-dependent dilation, sex differences in microvascular responsiveness to these stimuli in healthy young adults remain incompletely understood.

**Methods:** Cutaneous vasodilation was assessed in response to local heating to 39°C and 42°C and graded perfusion of ACh (10^-10^ to 10^-1^ mol/L) alone or concurrently with 15 mM N^G^-nitro-L-arginine methyl ester (L-NAME; NO synthase inhibitor) using laser-Doppler flowmetry coupled with intradermal microdialysis in 80 young adults (40 females).

**Results:** Local heating to 42°C elicited greater endothelium- and NO-dependent dilation than heating to 39°C in both groups (p<0.001), but no sex differences were observed at either temperature (p=0.65). ACh-induced endothelium-dependent dilation also was not different between sexes (p=0.08), but the NO-dependent component was greater in females than in males (p=0.01). In young females, menstrual cycle day (range: day 2-33) was not associated with endothelium- or NO-dependent dilation in response to any stimulus (all p≥0.19), regardless of hormonal contraceptive use.

**Conclusions:** Taken together, these findings suggest that sex differences in microvascular NO bioavailability in healthy young adults depend on the stimulus used to elicit cutaneous vasodilation and, in females, microvascular endothelium- and NO-dependent dilation are not meaningfully influenced by menstrual cycle phase.

## INTRODUCTION

The cutaneous microvasculature is an easily accessible microcirculatory bed for the minimally invasive investigation of vascular control mechanisms in humans.^1–3^ Indeed, both physiological (e.g., rapid local skin heating) and pharmacological (e.g., local administration of endothelium-dependent agonists) stimuli are commonly paired with intradermal microdialysis to mechanistically interrogate microvascular function in health and disease.^4–6^ Historically, local heating of the skin to 42°C has been the most frequently used technique to examine nitric oxide (NO)-mediated cutaneous vasodilation.^7–11^ In 2014, Choi *et al* demonstrated that a greater proportion of cutaneous vasodilation during local heating to 39°C is dependent on NO, proposing this temperature as a stimulus that more specifically isolates NO-mediated dilation and thereby enhances the sensitivity to detect improvements in microvascular endothelial function with acute or chronic interventions.^12^ This approach has since become more widely adopted, particularly in studies examining cutaneous NO bioavailability in healthy young adults.^4,13–16^ Acute localized administration of the endothelium-dependent agonist acetylcholine (ACh) to the cutaneous microvasculature, via intradermal microdialysis or iontophoresis, similarly probes the mechanistic regulation of endothelial function.^10,17–19^ For example, intradermal microdialysis perfusion of ACh, coupled with concurrent pharmacological NO synthase inhibition, is a routinely used approach for assessing microvascular NO-mediated endothelium-dependent dilation. However, despite the accessibility of these minimally invasive experimental techniques and their increasingly common use, whether there are sex-specific differences in cutaneous microvascular responsiveness to these stimuli in healthy young adults remains incompletely understood. This gap complicates both the interpretation and cross-study comparison of existing literature and makes it difficult to assess the rigor and reproducibility of studies proposing to use these approaches to understand either sex- or disease-related alterations in microvascular function.

Most studies using local heating to either 42°C or 39°C report no differences in cutaneous vasodilation between apparently healthy young males and females, though this is not a universal finding.^7,20–22^ The literature examining sex differences in the NO-dependent component of local heating-induced vasodilation is also conflicting, with studies providing evidence for reduced, greater, and equivalent NO-dependent dilation in males compared to females.^21–23^ While fewer studies have used ACh to pharmacologically probe sex differences in microvascular function in healthy young adults, these investigations also generally report no difference in ACh-induced vasodilation between sexes, although it has been reported that males may have a modestly greater NO-dependent component compared to females.^24–26^ The reasons for these inconsistencies remain unclear but may reflect differences in study design (e.g., consideration of menstrual cycle phase or hormonal contraceptive use in female participants), the absence of studies concurrently employing all three approaches within the same individuals, and the use of underpowered sample sizes.

Therefore, the purpose of this study was to assess cutaneous microvascular endothelium-dependent dilation, and the relative contribution of NO to this response, in a large sample of apparently healthy young males and females in response to rapid local heating to 42°C and 39°C and local intradermal microdialysis perfusion of ACh. We applied conventional analytical approaches frequently utilized by experts in the field, as well as novel methods detailed in our companion manuscript (Schwartz et al., 2026), to rigorously examine sex differences in all outcome measures. We hypothesized that endothelium-dependent dilation would not differ between sexes across all three stimuli, but that the NO-mediated component would be reduced in males in response to local heating and in females in response to ACh. To enhance ecological validity, both naturally-cycling females and those using hormonal contraceptives were enrolled and all females were tested without regard for menstrual cycle or hormonal contraceptive phase. However, consistent with recent guidelines for optimizing rigor and reproducibility when including premenopausal females,^27^ we also examined the potential influence of both hormonal contraceptive use and self-reported menstrual/hormonal contraceptive cycle day on endothelium- and NO-dependent dilation in response to each stimulus.

## METHODS

### Participants

Experimental protocols were approved by the Institutional Review Boards at the University of Delaware (2190901) and the University of Iowa (202404816). This study was registered on clinicaltrials.gov (unique identifier: NCT06499844). Verbal and written informed consent were voluntarily obtained from all participants prior to participation and according to the Declaration of Helsinki.

Eighty young adults (18-30 yrs; 40 males, 40 females) completed the study: 40 participants (20 males, 20 females) were tested at each study site, according to standard operating procedures jointly developed prior to study initiation. All participants completed a basic health screening that included a medical history and measurements of height, mass, and resting seated blood pressure and heart rate (SureSigns VS2+, Philips Healthcare, Andover, MA and Connex Spot Monitor, Welch Allyn, Skaneateles Falls, NY). Race and ethnicity were self-reported. Participants were excluded for any self-reported diagnosis of cardiometabolic disease or if body mass index was <18.5 or >35 kg/m^2^.

Twenty-one females reported current use of hormonal contraception (12 oral pill, 5 intrauterine device, 3 implant, 1 patch). The degree to which menstrual cycle or hormonal contraceptive phase influence cutaneous microvascular function remains equivocal.^28–30^ To improve generalizability, experimental visits were scheduled without regard to menstrual cycle phase. At the experimental visit, female participants self-reported the number of days since the start of their last menstrual period, and a urine pregnancy test confirmed the absence of pregnancy.

### Experimental Assessment of Microvascular Endothelium-Dependent Dilation

All participants completed a single laboratory visit to assess cutaneous microvascular endothelium-dependent vasodilation according to standard protocols extensively described by our team and others.^5–7,9,25^ All protocols were conducted in a quiet, thermoneutral environment with participants in a semi-recumbent position. Prior to the laboratory visit, participants were instructed to avoid caffeine for 12 hours and abstain from exercise and alcohol for 24 hours.

Using aseptic technique, four intradermal microdialysis fibers (CMA Linear 30 probe, 6 kDa; Harvard Apparatus, Holliston, MA, USA) were inserted into the dermal layer of the ventral forearm for local delivery of pharmacological agents. The use of pharmacological agents with intradermal microdialysis was approved by the U.S. Food and Drug Administration (IND no. 156,345). Pharmacological agents were prepared immediately before use, dissolved in lactated Ringer’s solution, filtered using sterile syringe microfilters (Acrodisc; Pall, Ann Arbor, MI, USA), and wrapped in foil to prevent degradation from light exposure. The protocols described below commenced after a ∼60-90 min hyperemia-resolution period, during which lactated Ringers or the site-specific pharmacological solutions were perfused through the microdialysis probes (2 μL/min; Bee Hive controller and Baby Bee microinfusion pump; BASi, West Lafayette, IN, USA). Cutaneous red blood cell flux was continually measured directly over each microdialysis probe via an integrated laser-Doppler flowmeter secured in a local heating unit set to a thermoneutral temperature (33°C; VP12 and VHP2; Moor Instruments, Wilmington, DE, USA). Automated brachial blood pressure was measured on the contralateral arm every 4-5 minutes throughout the protocol.

### Physiological Approach to Elicit Vasodilation: Local Heating

Following ∼10 minutes of baseline measurements at 33°C, endothelium-dependent dilation was assessed at two microdialysis sites using rapid local skin heating (0.5°C/sec) to either 39°C or 42°C.^4,6,7,9,12,21^ After the local heating-induced increase in red cell flux reached a stable plateau (∼40 mins), both sites were then perfused with N^G^-nitro-L-arginine methyl ester (L-NAME; 15mmol/L; Calbiochem) to non-selectively inhibit NO synthase and thus allow for the quantification of the relative contribution of NO to local-heating-induced endothelium-dependent dilation. After the subsequent post-L-NAME plateau in red cell flux was established (∼40 mins), the two sites were then perfused with the NO donor sodium nitroprusside (SNP; 28 mmol/L; United States Pharmacopeial, Rockville, MD, USA) and the temperature of the local heating units was simultaneously increased to 43°C to elicit maximal vasodilation.^21,25,31^

### Pharmacological Approach to Elicit Vasodilation: Acetylcholine Perfusion

Following ∼10 minutes of baseline measurements at 33°C, endothelium-dependent dilation was assessed in the remaining two microdialysis sites by perfusing ascending concentrations of ACh (10^-10^ to 10^-1^ mol/L; United States Pharmacopeia) alone (i.e., control) and during co-perfusion with L-NAME (15 mmol/L).^25,31^ Following the ACh dose-response protocol, maximal cutaneous vasodilation was elicited at each of these two sites as described above.

### Data and Statistical Analysis

Red blood cell flux was continuously recorded at 40 Hz (PowerLab and LabChart ADInstruments, Bella Vista, NSW, Australia). Cutaneous vascular conductance (CVC) was calculated as laser-Doppler flux (perfusion units) divided by mean arterial pressure and normalized as a percentage of the site-specific maximum (%CVC_max_). Although our companion paper employs a more comprehensive analytical approach to quantify endothelium- and NO-dependent dilation (Schwartz et al., 2026), the current analyses are restricted to those used in prior studies examining sex differences to facilitate direct comparison with the existing literature, with the exception of our novel method of analyzing ACh dose-response curves (described below).

For the local heating protocols, endothelium-dependent dilation was quantified as a stable 3–5-minute plateau in skin blood flow.^7,21,31^ NO-dependent dilation was calculated as a percentage of the local heating-induced plateau in cutaneous blood flow [(local heating plateau – L-NAME plateau) / (local heating plateau) x 100].^32^

For the ACh perfusion protocol, endothelium-dependent dilation was assessed from a stable 1-2 min plateau at the conclusion of each sequential dose of ACh.^25,31,33,34^ Because there is less of a consensus regarding the quantification of ACh-induced dilation, these data were analyzed as both the measured CVC at each dose and as the area under the curve (AUC; arbitrary units, a.u.) while accounting for blood flow during the thermoneutral baseline (GraphPad Prism 10.6.0, San Diego, CA). NO-dependent dilation was calculated as the difference in the AUC between sites.^25,34–36^ NO-dependent dilation was not derived if AUC at the L-NAME site was greater than AUC at the control site (n=6 males; n=7 females).

In addition, we applied the novel analytical approach extensively described in our companion paper (Schwartz et al., 2026), in which dose-response curves were fit using a four-parameter nonlinear mixed-effects model (Hill slope constrained to 1) with group-level fixed effects (top, bottom, logEC_50_) and subject-specific random effects (top, bottom, logEC_50_).^37^ This analytical approach enabled inclusion of all participants (n=80) at both control (ACh alone) and ACh+L-NAME sites, improving statistical power and reducing bias associated with traditional two-step modeling methods.^25,31,37^ The logEC_50_ (effective concentration resulting in 50% of the maximal response) was used as a secondary index of ACh-induced endothelium-dependent dilation. For completeness, the logEC_90_, the maximum response (top), and the basal or minimum response (bottom) were also derived.

Sample size was determined a priori (power=0.80, l1=0.05). Using previously published data from our laboratory with similar primary outcomes in response to local heating,^7^ we determined that n=11 per group (Cohen’s d=1.15) and n=40 per group (Cohen’s d=0.57) would provide ≥80% power to detect meaningful physiological differences in endothelium- and NO-dependent dilation between sexes, respectively. Given that menstrual cycle phase and hormonal contraceptive use were not experimentally controlled, we selected the more conservative estimate of n=40 per group as our enrollment target, which was achieved.

All data were assessed for normality via visual inspection of Q-Q plots, and sensitivity analysis confirmed results were robust to violations of homoscedasticity. Comparisons between groups for participant characteristics were made using unpaired Student’s t-tests and chi-square tests of independence for categorical variables. Group differences in endothelium- and NO-dependent dilation were analyzed using 2- or 3-way repeated measures ANOVA (sex x. temperature/site x. phase/dose) with Tukey post hoc corrections when appropriate (SAS 9.4, Cary, NC). Group differences in ACh-induced NO-dependent dilation were made using unpaired Student’s t-tests. In females, the associations between self-reported menstrual cycle day and endothelium- and NO-dependent dilation were assessed by multiple linear regression with hormonal contraceptive use included in the model as a categorial covariate (yes/no). Data are expressed as mean ± SD in tables and mean ± SEM in figures unless otherwise noted. Individual values are presented within each figure when appropriate. Significance was set at a < 0.05.

## RESULTS

Participant characteristics are presented in Table 1. The race and ethnicity distributions were not different between males and females (both p>0.05). There were no group differences in age (p=0.23). Both height and mass were greater in males (p < 0.001); however, body mass index did not differ between sexes (p=0.36). Seated systolic blood pressure was greater in males (p<0.001), but neither seated diastolic blood pressure nor heart rate differed between groups (both p≥0.05).

**Table 1.** Participant characteristics.

| Characteristic | Females | Males | p-value |
| --- | --- | --- | --- |
| Participants, *n* | 40 | 40 |  |
| Race, *n* (%) |  |  | 0.46 |
| White | 28 | 31 |  |
| Black | 4 | 1 |  |
| Asian | 5 | 7 |  |
| >1 Race | 2 | 1 |  |
| Unknown/not reported | 1 | 0 |  |
| Ethnicity, *n* (%) |  |  | 0.75 |
| Not Hispanic or Latino | 34 | 35 |  |
| Hispanic or Latino | 6 | 4 |  |
| Unknown/not reported | 0 | 1 |  |
| Age (yrs) | 22±3 | 22±3 | 0.23 |
| Height (cm) | 164.9±7.5 | 178.8±7.6 | < 0.00 |
| Mass (kg) | 66.1±10.6 | 80.4±14.4 | < 0.00 |
| BMI (kg/m<sup>2</sup>) | 24.3±3.5 | 25.0±3.5 | 0.36 |
| Seated systolic BP (mmHg) | 112±11 | 125±12 | < 0.001 |
| Seated diastolic BP mmHg) | 70±8 | 74±10 | 0.05 |
| Seated heart rate (bpm) | 76±10 | 73±11 | 0.18 |

Values are mean ± SD. BMI, body mass index; BP, blood pressure. Comparisons between groups were made using unpaired Student's t-tests and chi-square tests of independence for categorical variables.

There were no group differences in flux or absolute CVC at either baseline or at maximum between sites (Table 2; all group p≥0.26). Local heating to 42°C elicited greater endothelium-dependent dilation compared to 39°C in both groups, but there were no differences between females and males at either temperature (Fig.1A; group p=0.65). A greater proportion of endothelium-dependent dilation in response to local heating to 42°C was dependent on NO than that in response to 39°C in both females and males, but there also were no differences between groups at either temperature (Fig.1B; group p=0.74).

**Figure 1.**
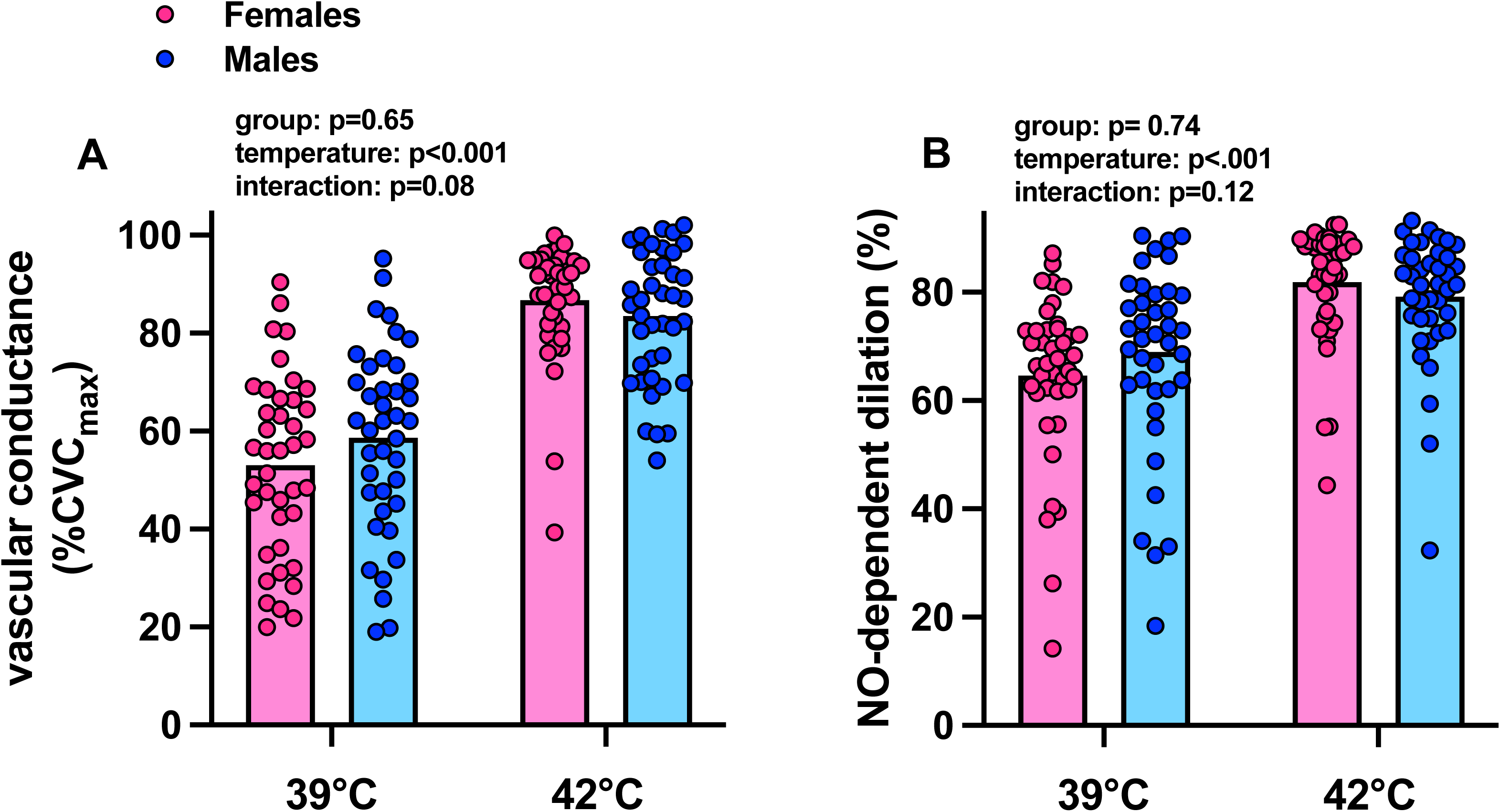
Local heating-induced cutaneous vasodilation. Vascular conductance in response to local heating to both 39°C and 42°C (panel A) and the relative portion mediated by nitric oxide (NO; panel B) in females (n=40; pink) and males (n=40; blue). Differences between sexes were compared using a two-way analysis of variance (group x. temperature) with post hoc comparisons performed using Tukey’s test if appropriate (all interaction p>0.08).

**Table 2.** Baseline and maximal flux and absolute cutaneous vascular conductance.

| Site | Flux (PU) |  | CVC (PU/mmHg) |  |
| --- | --- | --- | --- | --- |
|  | Baseline | Maximal | Baseline | Maximal |
| LH-39°C |  |  |  |  |
| Females | 23±23 | 170±90 | 0.31±0.29 | 2.39±1.37 |
| Males | 23±15 | 157±71 | 0.31±0.21 | 2.14±0.97 |
| LH-42°C |  |  |  |  |
| Females | 19±14 | 174±81 | 0.25±0.18 | 2.44±1.23 |
| Males | 17±15 | 139±71 | 0.22±0.18 | 1.83±0.73 |
| ACh |  |  |  |  |
| Females | 21±16 | 193±84 | 0.28±0.20 | 2.66±1.41 |
| Males | 22±14 | 194±87 | 0.29±0.18 | 2.64±1.38 |
| ACh+L-NAME |  |  |  |  |
| Females | 16±8 | 176±70 | 0.21±0.11 | 2.46±1.16 |
| Males | 18±13 | 180±93 | 0.24±0.17 | 2.45±1.29 |

Values are mean ± SD. Differences between females (n=40) and males (n=40) were compared using a three-way analysis of variance with post hoc comparisons performed using Tukey's test when appropriate (flux: group p=0.39, site p=0.092, phase p <0.091; CVC: group p=0.26, site p=0.02, phase p<0.01). PU, perfusion units (PU); CVC, cutaneous vascular conductance; LH, local heat; ACh, acetylcholine; L-NAME, N<sup>G</sup>-nitro-l-arginine methyl ester.

ACh elicited a dose-dependent increase in vascular conductance (Fig. 2A; dose p<0.01) that was blunted by concurrent L-NAME perfusion (Fig. 2A; site p<0.01) in both groups. In general, similar findings were obtained when the dose-response curves were modeled (Fig. 2B), indicated by a rightward shift in the logEC_50_ in the ACh+L-NAME site (Table 3, site p<0.01). Sex differences in ACh-induced dilation did not reach statistical significance (Fig. 2A, group p=0.08; Fig. 2B, group logEC_50_ p=0.12), although the top (maximum response) of the modeled curve was modestly greater in females than that in males (Table 3 and Fig. 2B, group p=0.04). A greater proportion of ACh-induced endothelium-dependent dilation was dependent on NO in females compared to males (Fig. 2C; p=0.012).

**Figure 2.**
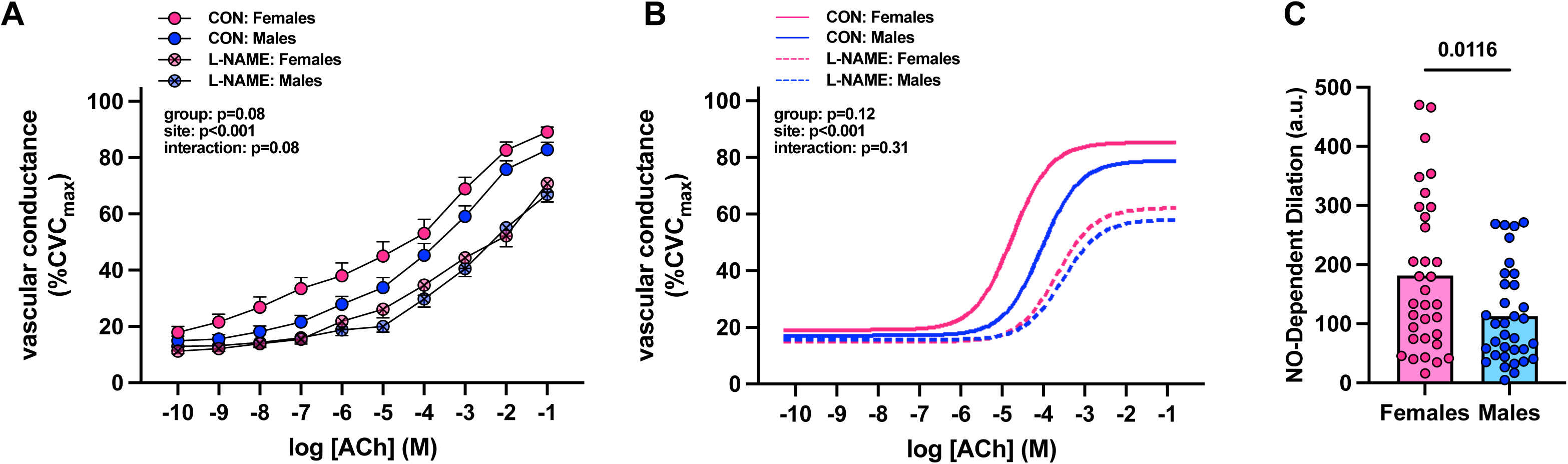
Acetylcholine (ACh)-induced cutaneous vasodilation. Vascular conductance in response to exogenous ACh perfusion alone (control site, CON; filled symbols/lines) and concurrent perfusion with N^G^-nitro-l-arginine methyl ester (L-NAME; hatched symbols/lines) at each concentration separately (panel A) and as a modeled sigmoidal dose-response curve (panel B) in females (n=40; pink) and males (n=40; blue). Differences between sexes were compared using a three-way (group x. site x. dose) or two-way analysis of variance (group x. logEC_50_), respectively, with post hoc comparisons performed using Tukey’s test if appropriate (all interaction p>0.08). The relative portion mediated by nitric oxide (NO; panel C) was calculated as the difference in the area under the curve (AUC; arbitrary units, a.u.) between control and L-NAME sites, while accounting for baseline blood flow at each (n=6 males, n=7 females). Differences between sexes were compared using unpaired Student’s t-tests.

**Table 3.** Group parameters for modeled acetylcholine dose-response curves.

| Curve Parameter | ACh |  | ACh+L-NAME |  |
| --- | --- | --- | --- | --- |
|  | Females | Males | Females | Males |
| Bottom | 19.41±1.4<br>[16.66, 22.15] | 17.4±1.34<br>[14.77, 20.03] | 15.16±1.33<br>[12.55, 17.78] | 15.85±.31<br>[13.28, 18.42] |
| Top | 87.44±.11<br>[83.3, 91.58] | 80.77±2.19 *<br>[76.49, 85.06] | 63.7±2.3<br>[59.19, 68.21] | 59.5±2.29 *<br>[55.0, 63.99] |
| EC<sub>50</sub> | -4.76±0.27<br>[-5.28, -4.24] | -4.09±0.27<br>[-4.62, -3.57] | -3.75±0.27<br>[-4.28, -3.21] | -3.61±0.28<br>[-4.15, -3.07] |
| EC<sub>90</sub> | -3.8±0.27<br>[-4.33, -3.28] | -3.14±0.27<br>[-3.66, -2.61] | -2.79±0.27<br>[-3.33, -2.26] | -2.65±0.28<br>[-3.2, -2.11] |

Values are means ± SE [95% confidence interval]. Differences between females (n=40) and males (n=40) were compared using a two-way analysis of variance for each parameter separately with post hoc comparisons performed using Tukey's test when appropriate. ACh, acetylcholine; L-NAME, N<sup>G</sup>-nitro-l-arginine methyl ester. \*p<0.05 vs females (at same site).

The range of self-reported day of menstruation at the time of the experimental visit was 2–33 days. Three females using intrauterine devices did not report menstrual cycles and were thus excluded from these analyses, as menstrual cycle day could not be determined. Menstrual cycle day was not associated with endothelium- or NO-dependent dilation in response to either physiological or pharmacological stimuli (Fig. 3A-D; all p≥0.19), even when controlling for hormonal contraceptive use.

**Figure 3.**
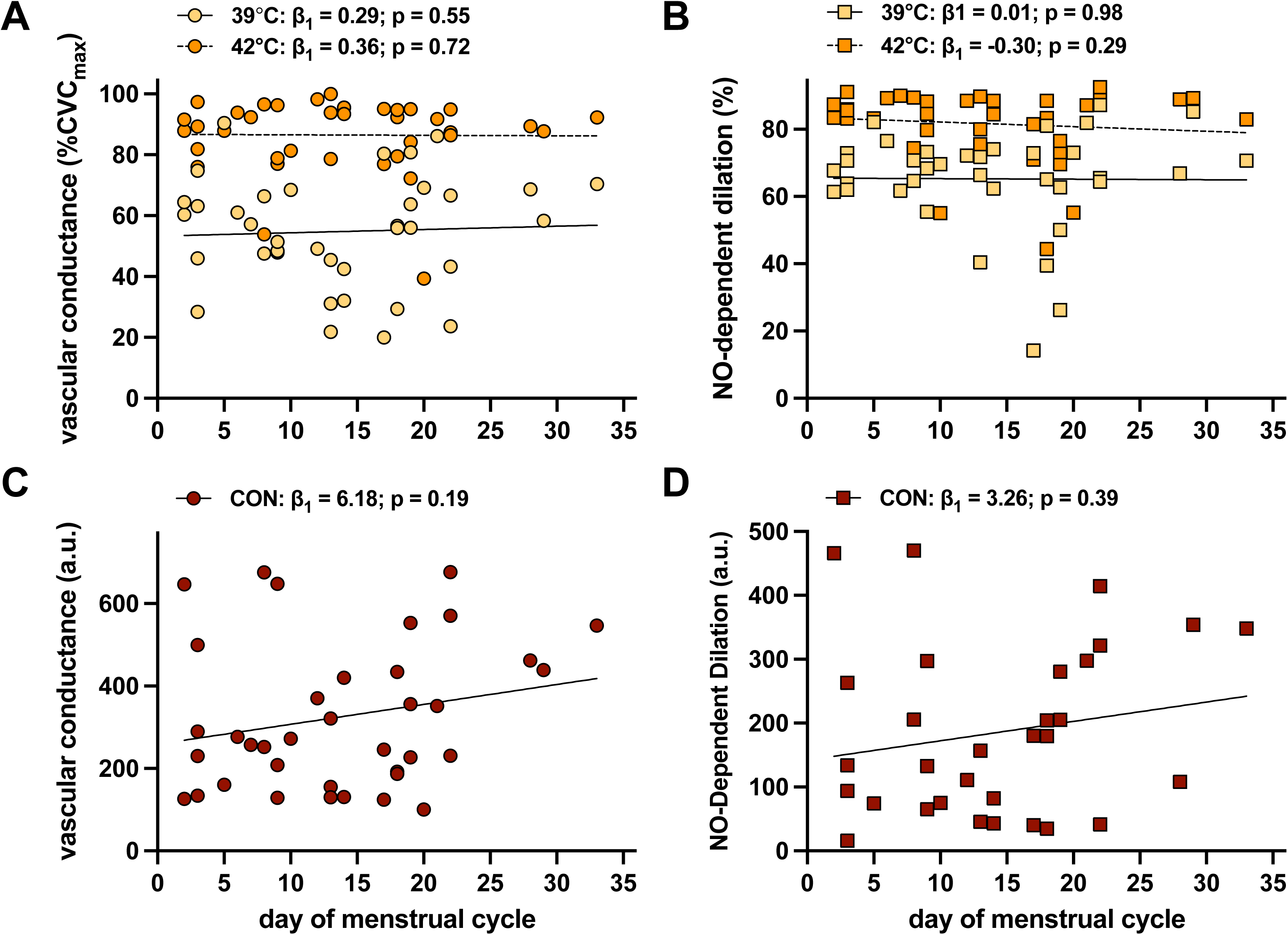
The association between self-reported day of the menstrual cycle and cutaneous vasodilation. Individual data and linear regressions of the relation between self-reported day of the menstrual cycle and both vascular conductance and nitric oxide (NO)-dependent dilation in response to local heating to 39°C (yellow symbols) and 42°C (orange symbols; panels A-B) and in response to exogenous acetylcholine (ACh) perfusion (panels C-D) in young females (n=40). For visual clarity, ACh-induced dilation is presented as area under the curve for the control site, and NO-dependent dilation was calculated as the difference in the area under the curve (AUC; arbitrary units, a.u.) between control and L-NAME sites, while accounting for baseline blood flow at each (n=31 females). In all models, hormonal contraceptive use was entered as a categorical covariate (yes/no).

## DISCUSSION

In the present study, we assessed sex differences in microvascular dilatory responses to rapid local heating to both 42°C and 39°C and local administration of exogenous ACh in a large sample of apparently healthy young males and females. The primary findings were, first, there were no differences between males and females in either local heating-induced endothelium- or NO-dependent dilation at either temperature. Second, we detected no sex differences in ACh-induced endothelium-dependent dilation, regardless of analytical approach. However, a larger proportion of ACh-induced dilation was dependent on NO in young females compared to young males. Lastly, in young females, there was no association between menstrual cycle day and endothelium- or NO-dependent dilation to either physiological or pharmacological stimulus, indicating that cutaneous microvascular function is not substantively altered across phases of the menstrual cycle. Taken together, these findings suggest that there are no sex differences in the cutaneous vasodilatory response to local heating or ACh perfusion in healthy young adults but sex differences in microvascular NO bioavailability depend on the stimulus. However, neither endothelium-dependent dilation nor the NO-mediated contribution is meaningfully influenced by menstrual cycle phase.

In both sexes, local heating to 42°C elicited greater endothelium-dependent dilation than heating to 39°C and, somewhat unexpectedly based on the literature,^12^ a larger NO-dependent component as well. Importantly, our data suggest that this temperature-dependent difference is not a sex-specific phenomenon, as the pattern was not different between groups. These findings are largely consistent with the majority of studies that have directly compared local-heating-induced vasodilation between young males and females, including prior work from our laboratories, nearly all of which report a lack of sex differences in the local heating-induced plateau in vascular conductance.^7,20–22^ However, this is not a universal finding, with one study reporting greater vasodilation in response to local heating to 39°C in young males compared to females.^23^ The reasons for this discrepancy are not clear, but given that we employed both heating stimuli simultaneously within the same participants, and with approximately double the sample size (n=40/group in the current study vs. n=7-18/group in prior work^20–23^), when taken together, our findings provide broad support for a lack of sex differences in the overall vasodilatory response to either 39°C or 42°C.

The existing literature is more equivocal regarding the NO-mediated component of local heating-induced vasodilation between sexes. In response to local heating to 42°C, there is some evidence for a ∼20% reduction in NO-dependent dilation in naturally-cycling young females not using hormonal contraceptives when tested during the early follicular phase of the menstrual cycle compared to males.^21^ In response to local heating to 39°C, some investigators report no differences in NO-dependent dilation between sexes, whereas others document a greater reliance on NO in young females.^22,23^ As above, the explanations for these conflicting findings are not readily apparent, though they are likely due, at least in part, to methodological differences between studies. For instance, Turner *et al* conducted subgroup analyses showing that NO-dependent dilation is greater in females during the placebo phase of oral contraceptive pill use compared to that in naturally-cycling females during the early follicular phase, which subsequently impacted the interpretation of sex differences in NO-dependent dilation when these females were considered as a single group compared to males.^23^

Surprisingly few studies have investigated sex differences in ACh-induced dilation in healthy adults, with the extant literature limited to secondary or tertiary comparisons within larger studies testing tangentially related hypotheses.^24–26^ Of these studies, all report no sex differences in endothelium-dependent dilation in response to intradermal microdialysis perfusion of ACh, either when administered sequentially across a range of physiologically-relevant concentrations or when delivered as single prolonged supraphysiological dose (2000 mM). Studies utilizing acetylcholine iontophoresis similarly report no differences between sexes in adults with underlying comorbidities.^38,39^ Consistent with this, we also show no differences in ACh-induced dilation alone or during concurrent NO synthase inhibition between young males and females. Similar results were obtained using our novel sigmoidal dose-response approach as a secondary index of vasodilatory responsiveness to ACh, as evidenced by the absence of group differences in the logEC_50_.

Despite the lack of sex differences in overall ACh-induced dilation, we detected a greater NO-dependent component in response to ACh in young females than in young males. These data may suggest that females rely to a greater extent on calcium-mediated activation of endothelial NO synthase, whereas males have a greater dependence on increased release of prostaglandin and/or endothelium-dependent hyperpolarizing factors to drive cutaneous vasodilation in response to ACh,^40,41^ though whether this reflects true sexual dimorphism in downstream signaling pathways or compensatory upregulation in males remains to be determined. Regardless, this finding was somewhat unexpected given the prior report that ACh-induced dilation elicited using the same experimental protocol as the current study is attenuated to a greater extent during concurrent NO synthase inhibition in young males compared to females, suggesting modestly greater NO-dependent dilation in young males, though this was observed in a relatively small sample (n=10/group).^25^ However, there is evidence for an enhanced reliance on NO-dependent mechanisms in response to ACh in mesenteric resistance arteries isolated from sexually mature female rats compared with those from males, mediated in part by a greater density and sensitivity of small- and intermediate-conductance calcium-activated potassium channels on the endothelium.^42^ These preclinical data are consistent with our current findings and highlight a potential underlying mechanism that warrants future investigation. Despite these divergent dominant pathways between sexes, the overall net magnitude of ACh-induced endothelium-dependent dilation appears comparable between young males and females, perhaps reflecting functional compensation across downstream signaling pathways.

To enhance ecological validity and real-world applicability, we did not experimentally control for either menstrual cycle phase or hormonal contraceptive pill phase during testing.^43,44^ Of the 40 female participants, approximately half (n=21) reported current use of hormonal contraceptives and half (n=19) did not. Experimental testing occurred over the full range of the menstrual cycle (days 2–33) with a largely uniform distribution. We observed a complete absence of any association between menstrual cycle day and either local heating- or ACh-induced endothelium- or NO-dependent dilation, even when statistically accounting for hormonal contraceptive use in the regression models. Although both estrogen and progesterone are vasoactive,^45–47^ these data argue against a physiologically detectable influence of menstrual cycle phase on cutaneous NO-mediated vasodilation in response to physiological or pharmacological stimuli in cross-sectional designs. Whether within-person cyclical variations in reproductive hormone profiles produce corresponding transient changes in microvascular function requires employing more intensive experimental designs in which females are tested repeatedly throughout a single menstrual cycle, ideally across several months. Collectively, these data suggest that endothelium-dependent dilation and its NO-mediated component are not systematically influenced by biological sex in healthy young adults, and that prior reports of sex differences in this context may reflect methodological heterogeneity and inadequate sample sizes rather than true physiological sexual dimorphism.

### Perspectives

In conclusion, our findings demonstrate that overall cutaneous endothelium-dependent dilation in response to commonly used physiological and pharmacological stimuli are not different between apparently healthy young males and females. There are also no sex differences in the NO-dependent component of vasodilation elicited by local heating to either 42°C or 39°C. However, the proportion of ACh-induced dilation mediated by NO-dependent mechanisms is greater in young females than in young males. Finally, menstrual cycle phase does not meaningfully impact endothelium- or NO-dependent dilation to either local heating or ACh perfusion. Using a large and appropriately powered sample, and with convergent results across both conventional and novel analytical approaches, these data address several notable inconsistencies in the existing literature examining sex differences in cutaneous microvascular function in healthy young adults and indicate that sexual dimorphism in NO bioavailability is dependent on the vasodilatory stimulus employed. Importantly, these conclusions were reached using a more ecologically valid and generalizable approach to including and testing premenopausal females that is consistent with consensus guidelines for best practices^27,48^—an approach we posit is most appropriate given the influence of reproductive hormones was not a primary research question. Collectively, these findings will inform more rigorous consideration of sex as a biological variable in studies designed to understand the mechanisms of disease- and risk-related microvascular dysfunction.

## Data Availability

All data produced in the present study are available upon reasonable request to the authors

## ACKNOWLEDGMENTS

We gratefully acknowledge the effort expended by the volunteer participants. We also thank Ruda Lee, PhD, Aaron Autler, MSW, Grace Maurer, MS, Joy Mochache, BS, and Navya Vadlamudi, BS for their technical assistance.

## SOURCES OF FUNDING

Research reported in this publication was supported by an Institutional Development Award (IDeA) from the National Institute of General Medical Sciences of the National Institutes of Health under award number 2P20GM113125.

## DISCLOSURES

The authors report no biomedical financial interests or potential conflicts of interest.

## NONSTANDARD ABBREVIATIONS

Ach: acetylcholine
AUC: area under the curve
CVC: cutaneous vascular conductance
L-NAME: N^G^-nitro-L-arginine methyl ester
NO: nitric oxide
SNP: sodium nitroprusside

## PATHOPHYSIOLOGICAL NOVELTY AND RELEVANCE

### What is new?

- There are no sex differences in local heating-induced endothelium- or NO-dependent dilation in young adults.
- Although there are no sex differences in ACh-induced dilation, the proportion dependent on NO is greater in females than males.
- In young females, menstrual cycle day is not associated with endothelium- or NO-dependent dilation to either local heating or ACh.

### What is relevant?

- Sex differences in microvascular NO bioavailability in healthy young adults depend on the vasodilatory stimulus but are not meaningfully influenced by menstrual cycle phase.

### Clinical/Pathophysiological Implications

- Collectively, these findings will inform more rigorous consideration of sex as a biological variable in studies designed to understand the mechanisms of disease- and risk-related microvascular dysfunction.

## Notes

### Competing Interest Statement

The authors have declared no competing interest.

### Clinical Trial

NCT06499844

### Author Declarations

Experimental protocols were approved by the Institutional Review Boards at the University of Delaware (2190901) and the University of Iowa (202404816).

